# Polygenic Risk Scores for Prediction of Breast Cancer in Korean women

**DOI:** 10.1101/2021.11.18.21266495

**Authors:** Yon Ho Jee, Weang-Kee Ho, Sohee Park, Douglas F. Easton, Soo-Hwang Teo, Keum Ji Jung, Peter Kraft

## Abstract

**Background:** Polygenic risk scores (PRSs) developed using European and Asian GWAS have been shown to have good discrimination in Asian women. However, prospective calibration of absolute risk prediction models based on a PRS or PRS combined with lifestyle clinical and environmental factors in Asian women is limited. We evaluate the discrimination and calibration of several breast cancer PRSs among Korean women; these PRSs were developed using Asian and/or European training samples and include between 11 and 947,621 variants.

**Methods:** For each PRS, we compared discrimination (area under the curve [AUC]) and calibration (expected-to-observed ratio [E/O]) of three absolute risk models among 41,031 women from the Korean Cancer Prevention Study (KCPS)-II Biobank: (i) a model using incidence, mortality, and risk factor distributions (reference inputs) among U.S. women and European relative risks; (ii) a recalibrated model, using Korean reference but European relative risks; and (iii) a fully Korean-based model using Korean reference and relative risk estimates from KCPS.

**Results:** All Asian and European PRS improved discrimination over lifestyle, clinical and environmental (Qx) factors in Korean women; a PRS trained using both European and Asian GWAS results led to the greatest improvement (Qx: AUC=0.65, Qx+PRS: AUC=0.72). U.S.- based absolute risk models overestimated the risks for women age ≥50 years, and this overestimation was larger for models that only included PRS (E/O=1.2 for women <50, E/O=2.7 for women ≥50). Recalibrated and Korean-based risk models had better calibration in the large, although the risk in the highest decile was consistently overestimated. Absolute risk projections suggest that risk-reducing lifestyle changes would lead to larger absolute risk reductions among women at higher PRS.

**Conclusions:** Incorporation of Asian and European PRS can improve discrimination in Korean women and may be useful for the risk-stratified interventions.

**Key Messages:**

⍰ Prospective validation of absolute risk prediction models combining lifestyle and polygenic risk scores in Asian women is limited.
⍰ We evaluated the calibration and discrimination of five PRSs developed using Asian and/or European training samples; two PRS were restricted to genome-wide significant SNPs, two included sub-genome-wide significant SNPs, and a multi-ancestry PRS using both European and Asian GWAS results.
⍰ Incorporation of PRS previously developed in Asian and European-ancestry populations can improve discrimination in Korean women.
⍰ Calibration improved for risk models that incorporate age-specific incidence rates from the target population relative to models that use external incidence rates
⍰ Our finding suggests that PRS may be useful for prioritizing individuals for targeted intervention on their lifestyle such as alcohol intake and obesity.
⍰ Further studies are needed to evaluate the value of incorporating PRS into risk models in ancestrally diverse populations.

## INTRODUCTION

Breast cancer is the leading cancer diagnosed among women in most countries in the world. While the incidence of breast cancer in Asian women is currently lower than that in Western countries, the age distribution of breast cancer incidence in Asian women is markedly different from that in the Western countries, with a peak at 45–49 years in the Asian countries vs. 60–70 years in the Western countries^1,2^. We previously found this age difference led to overestimation of risk in Korean women when conventional breast cancer risk models developed in European-ancestry populations were used^3^. This underscores the need to validate Western-derived risk prediction models in Asian women and adapt them to improve their predictive ability.

In addition to lifestyle, clinical and environmental breast cancer risk factors, genetic susceptibility can play an important role in the development of breast carcinogenesis^4^. A large proportion of genetic variation in risk for breast cancer is polygenic due to multiple common single nucleotide polymorphisms (SNPs) with a small risk individually. These common breast cancer susceptibility SNPs have been discovered by genome-wide association studies (GWAS). Most of these GWAS have been conducted in European-ancestry women^5–8^; those conducted in other populations—including Asian women^9–13^—have smaller cumulative sample sizes. The combined effects of these susceptibility SNPs can be summarized as polygenic risk scores (PRS) using training data from European-ancestry GWAS^14–16^, Asian GWAS, or both^17–19^. The incorporation of a 313-SNP PRS developed for European-ancestry women (69,732 controls and 88,916 cases) into classical risk prediction model improved discrimination and risk stratification in women of European descent^16^. A 46-SNP PRS developed in Asian women (22,113 controls and 22,013 cases) was shown to be less predictive than the European-derived 313-SNP PRS in Asian women^17^. The better performance of the European PRS than Asian PRS may be due to the larger sample size from which the European PRS was derived. This result held in Korean women (hazard ratio [HR] per unit standard deviation [SD]=1.57 for European PRS vs. HR per SD=1.40 for Asian PRS). However, few studies have assessed calibration of the European PRS and Asian PRS absolute risk models in Asian women. Moreover, prospective validation of absolute risk predictions from models incorporating both lifestyle and PRS in Asian women is limited.

We previously used Individualized Coherent Absolute Risk Estimation (iCARE)^20^ to validate three risk prediction models (the U.S.-based European-ancestry model, a recalibrated model, and a fully Korean-based model) based on classical breast cancer risk factors in a Korean population^21^. Here we evaluate the predictive capacity of five PRSs developed using Asian and/or European training samples; two PRS were restricted to genome-wide significant SNPs (PRS-11_ASN_ and PRS-136_EUR_), two included sub-genome-wide significant SNPs (PRS-42_ASN_ PRS-209_EUR_), and a combined PRS trained using both European and Asian GWAS results. We also assessed the improvement in risk prediction and risk stratification by incorporating the PRS into classical risk factor model.

## METHODS

### Study population for discrimination and calibration analyses

Externally-developed absolute risk models were evaluated in 41,031 women age 20-80 years at enrollment from the Korean Cancer Prevention Study-II (KCPS-II) Biobank. Study participants undertook routine health assessments at nationwide health promotion centers between 2004 and 2013. The study design and recruitment have been described in detail previously^21^. 705 breast cancer cases occurred over 10 years of follow-up. These 41,031 women were not a representative sample of the women with available DNA in the KCPS-II since cancer cases were oversampled for genotyping (Supplementary Figure 1). We account for this oversampling in calibration analyses using inverse-probability-of-sampling-weights as implemented in the iCARE software. Supplementary Table 1 shows the questionnaire risk factor distributions in the validation cohort. See Supplementary Methods and Supplementary Table 2 for more details on genotyping and study population for relative risk estimation. All participants gave written informed consent before participation. The Institutional Review Board of Yonsei University approved this study protocol (IRB approval number 4-2011-0277).

### Polygenic risk scores

We compared the performances of four PRSs developed using European-ancestry GWAS or Asian GWAS: (i) Asian genome-wide significant SNPs found in the Biobank Japan^13^, (ii) Asian sub-genome-wide significant SNPs trained using summary statistics from an Asian GWAS meta-analysis^17^, (iii) European genome-wide significant SNPs reported in the Breast Cancer Association Consortium (BCAC)^5^, and (iv) European sub-genome-wide significant SNPs included in European-based PRS^16^. We calculated PRS for breast cancer using the formula 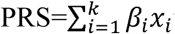 where *x*_*i*_ is the number of risk alleles (0,1,2) for SNP *i* and *β*_*i*_ is the corresponding weight. See Supplementary Table 3 and 4, Supplementary Figure 2, and Supplementary Methods for more details on SNP selection and weights used for each PRS. To compare the performance of these single-ancestry PRSs with a PRS trained using both European and Asian GWAS results, which was the best performing PRS in Ho et al.^17^, we included the results of discrimination and calibration for a multi-ancestry PRS (PRS_GW_EUR_ + PRS_GW_ASN_) derived using PRS-CSx method^22^. All PRSs were standardized to have mean 0 and SD 1 in the KCPS-II sample.

### Breast cancer absolute risk model validation and risk projections

Five-year absolute risks of breast cancer were calculated based on several external inputs: RR estimates for included risk factors; average age-specific absolute risk rates; the distribution of risk factors in the target population (estimated using a reference sample); and the age-specific competing mortality rates (Supplementary Methods). To evaluate the performance of five-year absolute risk models based on questionnaire data only^15^ (Qx), PRS only, and both questionnaire and PRS data (Qx+PRS), we used the iCARE software to estimate discrimination, measured by area under the curve (AUC), and calibration (overall expected-to-observed ratio [E/O] and expected versus observed incidence by expected risk deciles). We estimated cumulative and 10-year absolute risk trajectories across strata defined by genetic and modifiable risk profiles in the Korean-based Qx+PRS-CSx PRS model. We classified individuals in the top 20% of the PRS distribution as high PRS (corresponding RR≥1.33), those in the bottom 20% as low PRS (corresponding RR≤0.75), and those in the middle category (>20th to <80th percentile) as middle PRS. Individuals above the median of modifiable risk score distribution were classified as an elevated modifiable risk and those below the median were considered as reduced modifiable risk group (Supplementary Methods).

Descriptive statistics and regression analyses (Tables 1 and 2) were performed using SAS version 9.4 software (SAS Institute, Cary, NC). Absolute risks were evaluated with R version 4.0.3 software using the iCARE package 1.0.0.

**Table 1.** Mean and standard deviation of standardized polygenic risk scores with breast cancer risk

|  | Breast cancer events | No breast cancer |
| --- | --- | --- |
| n (%) | 705 (1.7) | 40,326 (98.3) |
| Mean (SD) of age at recruitment, in years | 44.09 (9.78) | 41.12 (11.41) |
| Mean (SD) of age of diagnosis, in years | 50.49 (10.03) | - |
| Mean (SD) of PRS-11<sub>ASN</sub> | 0.29 (1.02) | -0.01 (1) |
| Mean (SD) of PRS-42<sub>ASN</sub> | 0.33 (0.99) | -0.01 (1) |
| Mean (SD) of PRS-136<sub>EUR</sub> | 0.43 (1.02) | -0.01 (1) |
| Mean (SD) of PRS-209<sub>EUR</sub> | 0.43 (1.03) | -0.01 (1) |
| Mean (SD) of PRS<sub>GW_EUR</sub> + PRS<sub>GW_ASN</sub> | 0.53 (1.01) | 0.00 (1) |

SD=standard deviation; ASN=Asian; EUR=European.

**Table 2.** Association of polygenic risk scores and the occurrence of breast cancer.

| PRS discovery population | SNP selection | PRS | Number of SNPs published | Number of SNPs used in analyses | HR (95% CI)* | AUC (95% CI) |
| --- | --- | --- | --- | --- | --- | --- |
| Asian | Genome-wide significance<sup>1 13</sup> | PRS-11<sub>ASN</sub> | 11 | 11 | 1.35 (1.25 to 1.45) | 0.58 (0.56 to 0.61) |
| Asian | Sub-genome-wide significance<sup>2 17</sup> | PRS-42<sub>ASN</sub> | 46 | 42 | 1.40 (1.30 to 1.51) | 0.60 (0.58 to 0.62) |
| European | Genome-wide significance<sup>1 5</sup> | PRS-136<sub>EUR</sub> | 172 | 136 | 1.55 (1.44 to 1.66) | 0.62 (0.60 to 0.64) |
| European | Sub-genome-wide significance<sup>2 16</sup> | PRS-209<sub>EUR</sub> | 313 | 209 | 1.54 (1.43 to 1.66) | 0.62 (0.60 to 0.64) |
| Asian+European | Genome-wide significance<sup>2 17</sup> | PRS-CSx | 947,621 SNPs for PRS<sub>GW-EUR</sub><br>888,765 SNPs for PRS<sub>GW-ASN</sub> | 947,599 SNPs for PRS<sub>GW-EUR</sub><br>888,746 SNPs for PRS<sub>GW-ASN</sub> | 1.68 (1.57 to 1.81) | 0.65 (0.62 to 0.67) |

\*adjusted for principal component
<sup>1</sup>P-value < 5x10<sup>-8</sup>
<sup>2</sup>Clumping and threshold method
PRS = polygenic risk score; SNP = single-nucleotide polymorphism; HR = hazard ratio; AUC = area under the curve; CI = confidence interval; ASN=Asian; EUR=European.

## RESULTS

### Evaluation of PRSs in KPCS-II Biobank

Four single-ancestry PRSs and one multi-ancestry PRS were constructed using previously reported SNPs that passed imputation R^2^>0.8 in the KCPS-II: i) Asian genome-wide significant SNPs (PRS-11_ASN_); ii) Asian polygenic SNPs (PRS-42_ASN_); iii) European genome-wide significant SNPs (PRS-136_EUR_); iv) European polygenic SNPs (PRS-209_EUR_); and v) PRS-CSx method (PRS_GW_EUR_ + PRS_GW_ASN_). All PRSs had higher mean in cases than controls (Table 1). Among breast cancer cases, the mean PRS was higher for the European-based PRS than for the Asian-based PRS (PRS-11_ASN_: 0.29 vs. PRS-136_EUR_: 0.43, PRS-42_ASN_: 0.33 vs. PRS-209_EUR_: 0.43) and highest for PRS-CSx (0.53).

Table 2 shows the estimated HR per unit increase of PRS and AUC for breast cancer. Compared with the Asian PRS, the European PRS had larger effect sizes (PRS-209_EUR_: HR per SD = 1.54 vs. PRS-42_ASN_: HR per SD = 1.40) and a greater discrimination (PRS-209_EUR_: AUC = 0.62 vs. PRS-42_ASN_: AUC = 0.60) in the KCPS-II. For Asian PRS, PRS-42_ASN_ had a larger estimated HR and AUC than PRS-11_ASN_ (PRS-11_ASN_: HR per SD = 1.35, 95% confidence interval (CI) = 1.25, 1.45, AUC = 0.58). On the other hand, there was little difference in HR and AUC between PRS based on European genome-wide significant SNPs and sub-genome-wide significant SNPs (PRS-136_EUR_: HR per SD = 1.55, 95% CI = 1.44, 1.66, AUC = 0.62; PRS-209_EUR_: HR per SD = 1.54, 95% CI = 1.43, 1.66, AUC = 0.62). PRS generated using PRS-CSx showed the strongest association with breast cancer risk (PRS-CSx: HR per SD = 1.68, 95% CI = 1.57, 1.81, AUC = 0.65).

We then evaluated the predictive performance of models incorporating the PRS into absolute risk models along with conventional questionnaire-based risk factors (Qx). The incorporation of PRS improved discrimination overall (Figure 1A, Supplementary Figure 3, Supplementary Table 5 and 6). PRS-CSx showed the largest improvement when incorporated with questionnaire-based risk factors (Qx: AUC=0.65, Qx+PRS-CSx: AUC=0.72 among women age <50, Qx: AUC=0.54, Qx+PRS-CSx: AUC=0.63 among women age 50+ in Korean-based model). The improvement was slightly greater for the incorporation of European PRS compared to the incorporation of Asian PRS, especially among women of age 50+ (Qx: AUC=0.54, Qx+PRS-42_ASN_: AUC=0.59, Qx+PRS-209_EUR_: AUC=0.60 in Korean-based model). For Asian PRS, the combined model with PRS-42_ASN_ had a slightly larger AUC than PRS-11_ASN_ (Korean-based model: Qx+PRS-11_ASN_ =0.66 vs. Qx+PRS-42_ASN_ =0.68 in age <50; Qx+PRS-11_ASN_ =0.58 vs. Qx+PRS-42_ASN_ =0.59 in age 50+). Such difference was minimal between combined model with PRS-136_EUR_ and PRS-209_EUR_.

**Figure 1.**
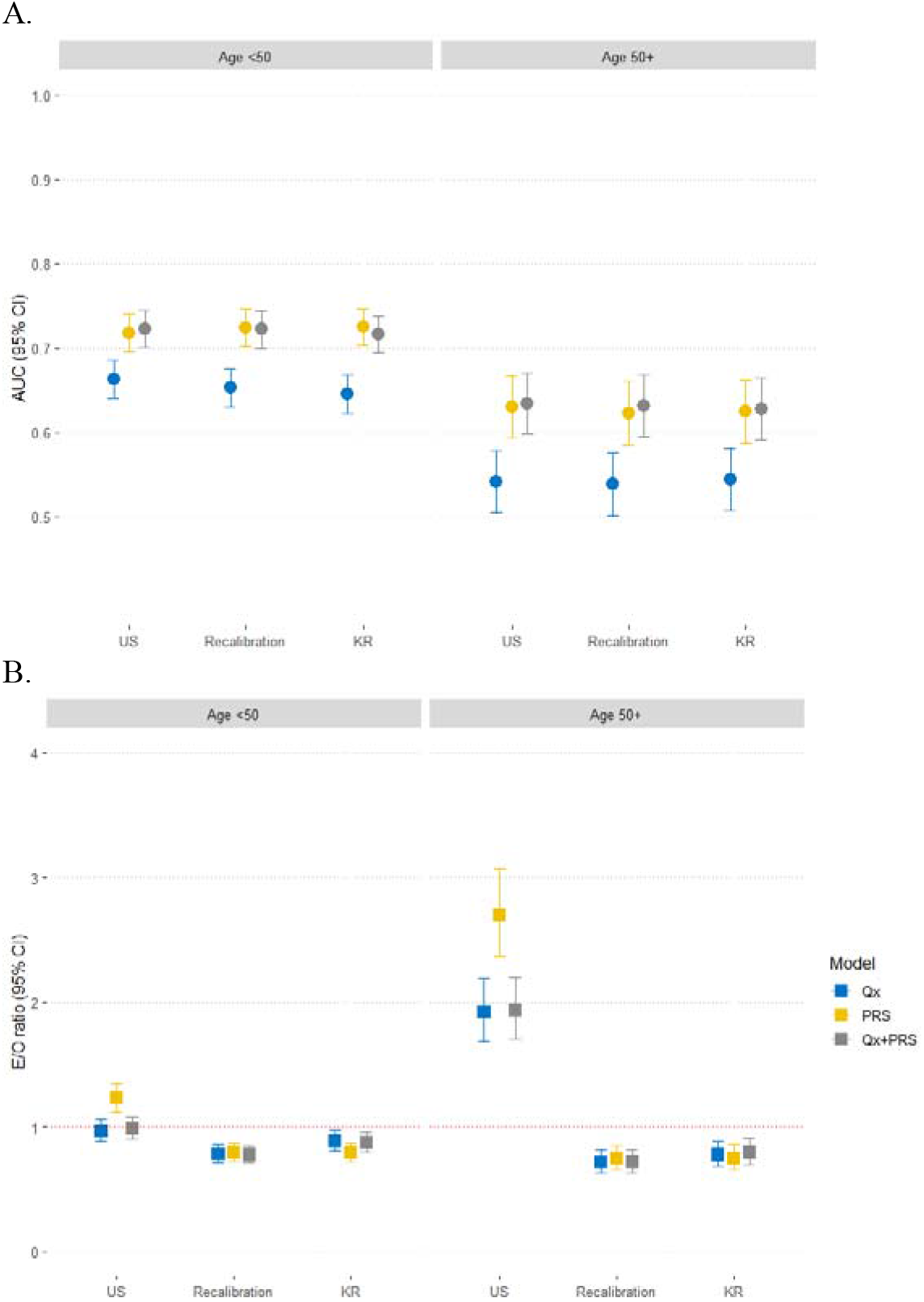
Discrimination and calibration of PRS-CSx for the breast cancer risk prediction models validated. (A) Discrimination and (B) calibration. Qx: questionnaire, US: a U.S.-based European-ancestry model, using incidence, mortality, and risk factor distributions among U.S. non-Hispanic white women and European-ancestry relative risks; Recalibration: a recalibrated model, using Korean incidence/ mortality and risk factor distributions but European-ancestry relative risks; and KR: a fully Korean-based model using Korean incidence/ mortality and risk factor distributions and relative risks estimates from the Korean Cancer Prevention Study. Area under the curve (AUC) and Expected/Observed (E/O) ratio estimates across first and second 5-years of follow-up periods were calculated using the fixed effects inverse variance weighting method, excluding women diagnosed with breast cancer or lost to follow-up in the first 5 years from the second 5 years of follow up.

Model calibration differed depending on the population incidence rates and reference population used to calculate absolute risks and whether PRS and/or questionnaire risk factor data were included (Figure 1B, Supplementary Figure 4, Supplementary Table 5 and 7). We found that PRS-only models using U.S.-based incidence rates overestimated the risks for all women, but particularly those older than 50: for all PRSs, the E/O ratio was 1.2 for women younger than 50 and 2.7 for women older than 50. In contrast, models incorporating questionnaire risk factor data only or both PRS and questionnaire data slightly underestimated risk among women younger than 50 (E/O ranging from 0.97 to 0.99) and modestly overestimated risk for women over 50 (E/O=1.9). Further recalibrations of the models using the Korean incidence and mortality rates, risk factor distributions from Korean population, and RR estimates from Korean population showed improved calibration, especially among older women. The change in calibration was particularly clear for the PRS-only models, especially among women over 50 [E/O=0.75 for the recalibrated and 0.79 for the Korean-based models].

Figure 2 presents calibration plots for the absolute risk model based on Qx factors, PRS-CSx and Korean incidence and mortality rates, risk factor distributions and relative risks. The decile-specific expected risks are largely linearly related to the observed risks. Consistent with calibration-in-the-large results, observed risks for the first nine deciles are mostly slightly larger than the expected risks. However, the model overestimates risks for women in the tenth decile (E/O=1.10 for women younger than 50 and 1.13 for women over 50).

**Figure 2.**
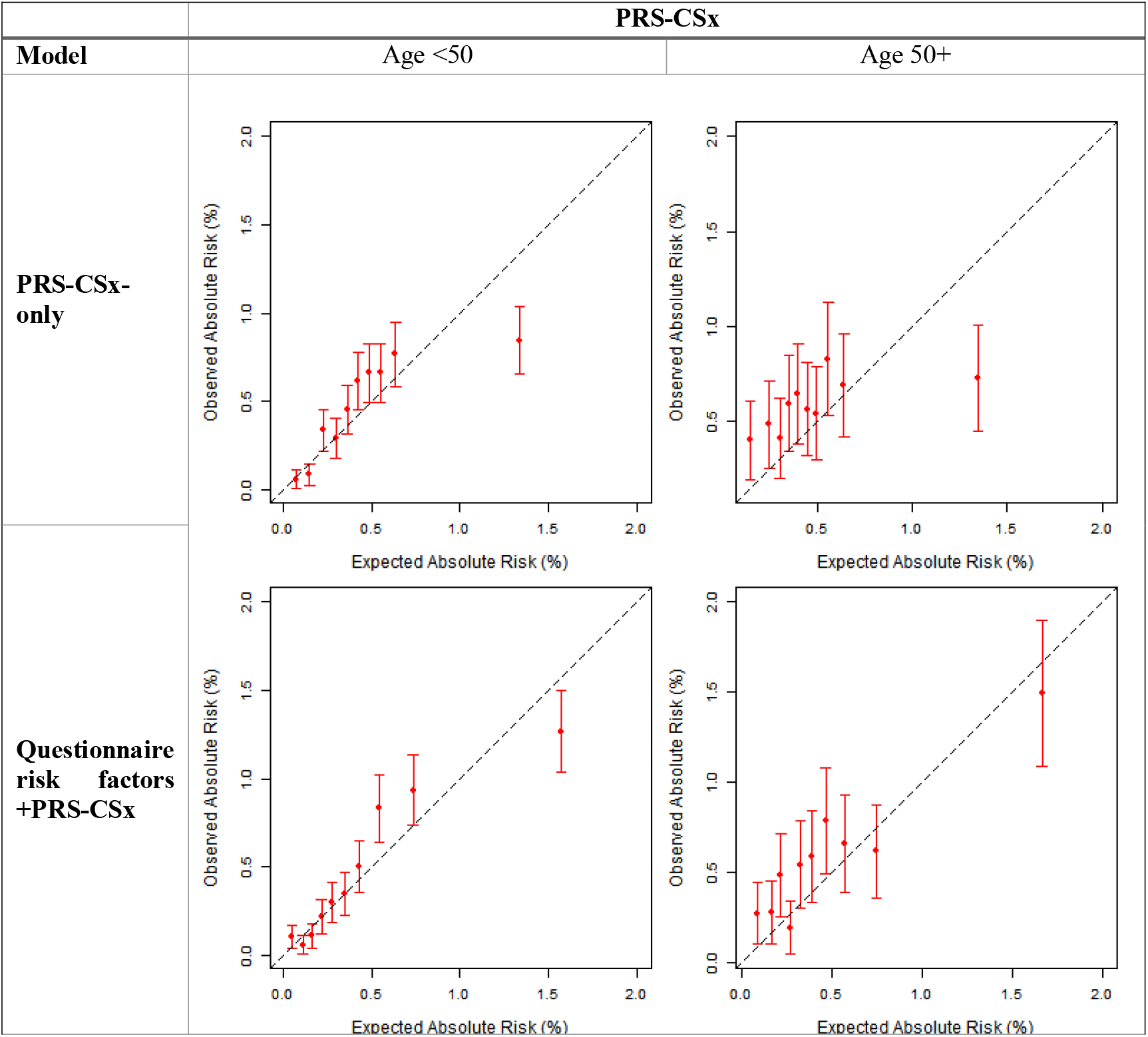
Absolute risk calibration for breast cancer risk prediction models in the Korean Cancer Prevention Study-II Biobank using PRS-CSx (Korean-based model*) *Korean-based model using Korean incidence mortality and risk-factor distributions and relative risk estimates from the Korean Cancer Prevention Study. For each decile, observed absolute risks and expected absolute risks were meta-analyzed between the first and second 5-years of follow-up using inverse variance weighting method.

### Absolute breast cancer risk predictions

We explored cumulative and 10-year absolute risk trajectories across strata defined by PRS-CSx and modifiable risk profiles in Korean-based model (Figure 3). The risk of developing breast cancer by age 80 was 1.2% (bootstrap 95% CI=0.6%-2.7%) for women in the low PRS group with reduced modifiable risk and 10.3% (bootstrap 95% CI=3.4%-14.2%) for those in the high PRS group with elevated modifiable risk (Supplementary Table 8). This model suggests that interventions on modifiable risk factors have the potential to reduce breast cancer risk, even among women at high risk due to their inherited genetics. Moreover, the differences in absolute risk between women with elevated and reduced modifiable risks were larger for women with higher PRS. The amount of risk difference between elevated and reduced modifiable risk was higher in higher PRS group (∼5% in the high PRS, ∼3% in the middle PRS, and ∼1% in the low PRS group).

**Figure 3.**
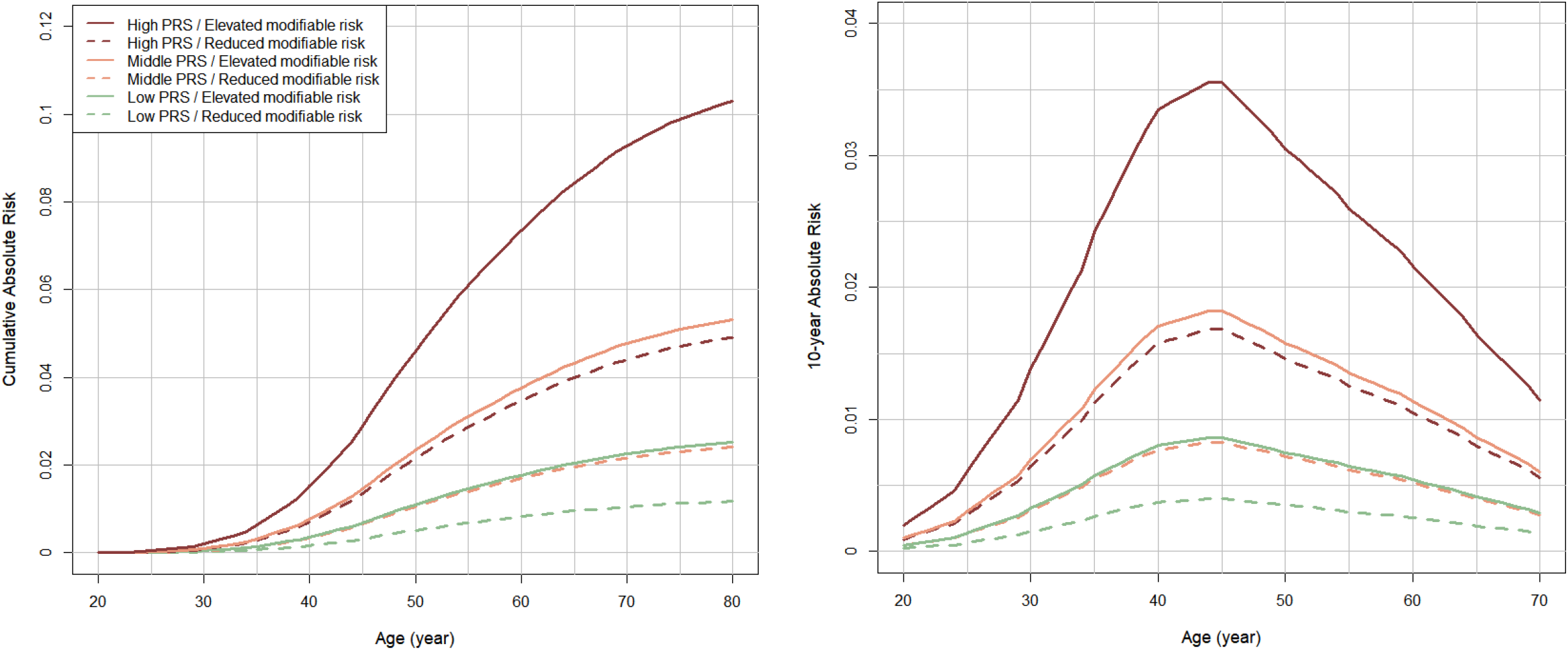
Lifetime and 10-year risk trajectories across strata defined by modifiable risk factors and percentiles of the PRS distribution using the combined PRS-CSx results (from the Korean model) Modifiable risk factors include body-mass index, oral contraceptive use, alcohol intake, and hormonal replacement therapy use. (High PRS: ≥80^th^; Middle PRS: 20^th^-80^th^; Low PRS: ≤20^th^; Elevated modifiable risk: ≥median; Reduced modifiable risk: <median)

## DISCUSSION

In this study evaluating five PRSs for breast cancer risk prediction in Korean women, we found that a PRS developed using both European-ancestry and Asian GWAS had better discrimination than PRS developed using European-ancestry or Asian GWAS alone. We also observed that inclusion of polygenic variants in additional to classical risk factors improved discrimination in Korean women. The calibration of absolute risk models depended on the source of information on average age-specific incidence rates, competing mortality, and risk factor distributions. Models that used age-specific incidence from an external population had poorer calibration than models using incidence rates from the target population; models derived using U.S. age-specific incidence rates showed poor calibration in the KCPS-II (E/O from 1.9 to 2.7 among women older than 50). This miscalibration was largest for risk models that only included PRS. Models using Korean incidence rates had good calibration in the large (E/O from 0.72 to 0.89), and estimated risks were correlated with observed risks, with the exception of the highest predicted risk decile, where risks tended to be overestimated. Our absolute risk projections suggest that a larger absolute risk reduction would occur among women at higher PRS by shifting to a healthier lifestyle.

We found that current European PRS showed better discrimination than current Asian PRS in a Korean population. A previous study also reported lower effect sizes and poorer performance of a Asian PRS (ORs per 1 SD: 1.10∼1.41 and corresponding AUCs: 0.533∼0.586) than those for a European-ancestry based 287-SNP PRS derived from 313-SNP PRS (odds ratio [OR] per 1 SD=1.51, AUC=0.617)^23^. Similarly, discrimination of European PRS (PRS-209_EUR_) in this study was slightly lower in Korean women (AUC=0.62, HR per SD=1.54) than that in European-ancestry women (AUC=0.63, OR per SD=1.61)^16^. We observed that a multi-ancestry PRS using both European and Asian GWAS results performed better than all four single-ancestry PRSs, which was in line with previous finding^17^. These findings highlight the challenges in transportability of current PRS across different populations, which arise from the overwhelming abundance of European-descent studies and the dearth of well-powered studies in diverse populations. Factors such as allele frequencies, linkage disequilibrium patterns, and demographic history and natural selection different across populations could lead to such differential transferability^24,25^. Hence, an Asian PRS with as large training sample size as European PRS may perform better among Korean women. It should be also noted that other existing PRS methods such as Stacked Clumping and Thresholding^26^ and LDpred2^27^ may potentially improve the performance significantly compared to genome-wide and sub-genome-wide significant SNPs PRS.

The U.S.-based absolute risk models were well calibrated among Korean women <50 years but overestimated the risks for those age ≥50 years, even after the incorporation of PRS. Notably, the overestimation was more extreme in all U.S.-based PRS-only models. This happens because the E/O estimate in the U.S.-based PRS-only model is completely driven by the differences in average incidence between the U.S. and KCPS-II, whereas the E/O estimates for the U.S.-based Qx-only and Qx+PRS models are driven by both the difference in average incidence and the difference in risk factor distributions. It turns out the latter two differences cancel out, making the calibration for Qx-only and Qx+PRS models look better than PRS-only models. The overestimation was corrected by recalibrating the models using the Korean-specific inputs, which underscores the importance of tailoring absolute risk models to the target population.

Consistent with previous studies^14,30^, our analysis indicates that a larger absolute risk reduction would occur among women at higher PRS by improving their lifestyle. This suggests that focusing on high-risk individuals could yield higher benefits of preventing cancers for certain risk factor modification interventions that may not be applicable to the whole population due to cost and other considerations. Given that young Asian women today are experiencing a dramatic increase in breast cancer incidence^1^, PRS could be used to adjust the optimal age for screening initiation and/or intensity to maximize the early detection of aggressive cancers, while minimizing the harms of screening in Asian women. For example, based on the distribution of risk factors at baseline in the KCPS-II (who were born between 1924 and 1993), the proportion of Korean women who had 10-year risk over 2.3% at age 40 was 10% in Qx+PRS model and 8% in PRS model. Similarly, a prior study projected that, based on their PRS, 12% of Chinese women born between 1960 and 1969 had 10-year risk over 2.3% at age 40—this being a recommended risk threshold for screening initiation— whereas the proportion of women born after 1979 passing that threshold at age 40 would rise to 29%^17^. However, ultimately evidence from clinical trials will be needed to understand the true effect of an intervention for the underlying population. Three trials of personalized risk-based breast screening incorporating PRS are underway to evaluate the efficacy, safety, and acceptability of risk-based screening in the USA and Europe^31,32^. Such clinical evaluation of PRS is urgently needed in Asian populations, where the burden of breast cancer is growing due to its dramatic increase in breast cancer incidence.

There are some limitations in our study. First, we could not consider subtypes of breast cancer since data were not available in the KCPS-II Biobank. Given the distributions of breast cancer subtypes are different between Asia and Western countries^33^, the prediction capacity may differ by subtypes in different populations. Second, although we provide important insight on the predictive capacity of multiple PRSs in a Korean population, our findings may not be generalizable to other Asian countries. Furthermore, since PRS-11_ASN_ was entirely based on BBJ, the PRS may be suboptimal for valid estimation of breast cancer risk in Korean women. Ho et al. reported that the distribution of this PRS was different across seven Asian countries, with the magnitude of differences consistent with genetic distance between these ethnic groups, confirming the importance of ethnic-specific calibration for valid estimation of breast cancer risk^23^. Third, we only used baseline information for first and second 5-years of follow-up and did not consider the changes in the risk factors during the follow-up period. Although this may have caused measurement error, we anticipate the magnitude of the error would not be large for most of the variables such as age at menarche, age at first birth, and family history of breast cancer. Fourth, although we had information of a large number of risk factors, we lacked data on several known risk factors. For instance, breastfeeding has been found to be the strongest protective factor in Korean women, whereas in European-ancestry women, the protective effect is relatively small^34,35^. In addition, Asian women have been reported to have denser breasts on mammography, which could increase their breast cancer risk^36,37^. Further validation studies using more comprehensive and Asian-specific risk factor models, along with Asian-specific PRS are needed. Finally, our results are based on observed associations between modifiable risk factors and breast cancer risk, not randomized trials of risk-factor-modifying interventions (e.g. guided dietary and physical activity changes). Consequently, our results cannot be interpreted as the expected risk reductions from risk-factor modifications without making additional assumptions (no unmeasured confounding, consistency of the interventions). Further research would be needed to establish the effect of risk-stratified interventions; we include these observational results to illustrate the potential impact of such a strategy.

Our study provides a comprehensive description of the utility of genetic and modifiable risk factors for the breast cancer risk prediction in a Korean population-based cohort. We established absolute risk models to reflect the age-specific incidence rates, distribution of risk factors, and RRs in the U.S. and Korea. Moreover, we evaluated model calibration stratified by levels of risk, which can be useful for risk-based breast cancer prevention and screening by identifying individuals at the extremes of risk.

## CONCLUSION

We have shown that incorporation of PRS previously developed in Asian and European-ancestry populations can improve discrimination in Korean women. Our findings suggest that PRS may be useful for motivating targeted prevention in high PRS group before they accumulate a high burden of modifiable risk factors. Larger Asian training samples should improve PRS discrimination among Korean women. Further studies are needed to evaluate the value of incorporating additional information on factors into a model in ancestrally diverse populations.

## Data Availability

All data produced in the present study are available upon reasonable request to the authors.

## NOTES

The funders had no role in the design of the study; the collection, analysis, and interpretation of the data; the writing of the manuscript; and the decision to submit the manuscript for publication.

Conceptualization: YHJ, PK; Data curation: KKJ; Formal analysis: YHJ, PK; Resources: KKJ, WKH, SHT; Supervision: PK; Writing – original draft: YHJ, PK; Writing – review & editing: all authors.

The authors have no conflicts of interest related to this work.

