## Supplementary Material for "Polygenic Risk Scores for Prediction of Breast Cancer in Korean women"

**SUPPLEMANTRY MATERIALS**

### **Supplementary Methods**

#### Study Population for Validation

*Data collection*

Data on reproductive and lifestyle factors were collected at enrollment: age at menarche, age at menopause, parity, age at first birth, oral contraceptive (OC) use (never, ever), hormone replacement therapy (HRT) use, alcohol intake, history of benign breast disease (BBD), and family history of breast cancer. Height and weight were measured while participants wore light clothing. Body mass index (BMI) was calculated as the weight (kg) divided by the height squared (m^2^).

*Genotyping methods.*

All samples in the validation cohort were genotyped using Illumina Global Screening Array v2.0. Based on a subset of autosomal variants with minor allele frequency (MAF) ≥ 0.01 and genotype call rate ≥ 98%, samples with call rate < 98% and extremes in heterozygosity were excluded. Data were imputed using SHAPEIT [1] for phasing and MINIMAC3 [2] for imputation, with 1000 Genomes Project (Phase 3) as the reference panel [3]. Post-imputation quality was evaluated using the R^2^ as provided by MIMINAC3 [2]. We excluded SNPs with R^2^ below 0.80 which were taken to be indicative of poor imputation quality and were not analyzed. After the exclusion, principal components were calculated using the imputed data that passed quality control using PLINK. The number of principal components needed to identify genetic outliers and define ancestry-informative covariates was determined based on a scree plot.

*Follow-up for breast cancer.*

The outcome was incidence of breast cancer (ICD-10 codes C50). The follow-up was nearly 100% complete because all participants have a unique identification number assigned at birth, allowing linkage with the national cancer registry and hospital admission records. The data on cancer diagnoses were obtained based on histologic type, resulting in high accuracy.

#### Study Population for Relative Risk Estimation

The Korean Cancer Prevention Study (KCPS) was used to independently estimate the relative risks (RRs) for breast cancer risk factors (Supplementary Table 2). The KCPS is a 1.3-million-member prospective cohort study, designed to assess risk factors for mortality, incidence, and hospital admission from cancer, with a follow-up of 25 years [4]. The KCPS cohort includes the 443,627 women ages 20–80 years who received health insurance from the Korean Medical Insurance Corporation and who had biennial medical evaluations between 1992 and 1995. The collection of risk factors was similarly done to the KCPS-II. Because history of BBD was not asked for women in the KCPS, we defined history of BBD based on ICD-10 code D24. In the KCPS cohort, an incident breast cancer was coded based on a hospital admission for a cancer diagnosis.

#### PRS analysis

*SNPs selection for each PRS*

We removed SNPs with low imputation quality (R^2^ < 0.8) in the KCPS-II. There was one SNP overlapping between the Asian genome-wide significant SNPs and the sub-genome-wide significant SNPs and 40 SNPs overlapping between European genome-wide significant SNPs and sub-genome-wide significant SNPs. For the PRS based on genome-wide significant SNPs, the genome-wide significance threshold was set at p < 5 × 10^−8^, and the sub-genome-wide-significant PRS were developed by clumping and thresholding (C+T) methods [5, 6]. The Asian genome-wide significant SNPs included significant SNPs at p < 5 × 10^−8^ after clumping (R^2^=0.1) using the East Asian 1000 Genomes reference panel. The European genome-wide significant SNPs were drawn from collaborations of multiple studies including the Breast Cancer Association Consortium (BCAC). Summary statistics for all SNPs used in derivation of four ancestry specific PRSs are provided in Supplementary Table 4. Weights used in the calculation of PRS-GW_ASN_ and PRS-GW_EUR_ are available in Supplementary Tables 3 and 4 of Ho et al. [6]. PRS-CSx is constructed by taking a weighted average: PRS-CSx =0.54881 + 0.16856 PRS_GW_ASN_ + 0.38484 PRS_GW_EUR_ [6].

*Weights used for each PRS*

For the PRS based on genome-wide significant SNPs, we used publicly available weights from a GWAS of Japanese or European-ancestry women [7, 8]. For the Asian and European sub-genome-wide significant PRS, we used the corresponding published weights [5, 6]. All PRS were standardized to have mean zero and standard deviation (SD) one in the total sample.

*Association analyses*

A Cox proportional hazards regression was used to estimate HRs per SD of each PRS (time scale = number of years since baseline, adjusted for the first 2 principal components). We calculated area under the receiver operating characteristic curve (AUC) using logistic regression.

#### Validation of Breast Cancer Risk Models

The questionnaire-based models included baseline data on reproductive, anthropometric, behavioral, and clinical risk factors: age at menarche, age at menopause, parity, age at first birth, oral contraceptive use, hormone replacement therapy use, BMI, height, alcohol intake, history of benign breast disease, and family history of breast cancer.

For each RR model incorporating questionnaire data, we evaluated three absolute risk models [9] in the KCPS-II Biobank: (i) a U.S.-based European-ancestry model, using incidence, mortality, and risk factor distributions among U.S. non-Hispanic white women and European-ancestry RRs; (ii) a recalibrated model, using Korean incidence/ mortality and risk factor distributions but European-ancestry RRs; and (iii) a fully Korean-based model using Korean incidence/ mortality and risk factor distributions and RR estimates from the KCPS. For the RR associated with a 1-SD difference in PRS, we used external estimates in models (i) and (ii) (from European-ancestry studies for European-ancestry PRS and from Asian-ancestry studies for Asian-ancestry PRS) and estimates from the KCPS-II for model (iii). Supplementary Table 2 provides detailed descriptions of RR estimates included in the models and population distribution.

iCARE calculates five-year absolute risks based on several external inputs: RR estimates for included risk factors; average age-specific absolute risk rates; the distribution of risk factors in the target population (estimated using a reference sample); and the age-specific competing mortality rates. iCARE then compares the predicted absolute risks to the observed incidence. We performed these discrimination and calibration analyses separately for the first and second 5-years of follow-up, excluding women diagnosed with breast cancer or lost to follow-up in the first 5 years from the second 5 years of follow up. Summary AUC and E/O estimates across these two follow-up periods were then calculated using the fixed effects inverse variance weighting method.

#### U.S. and Korean Reference Dataset

To obtain information on risk factor distributions, iCARE uses an additional individual-level reference dataset of risk factors representing each population. The reference datasets were 2010 National Health and Nutrition Examination Survey (NHANES) for the U.S.-based model and 2010-2012 Korean NHANES (KNHANES) for the recalibrated model and the Korean-based model. Details on classical risk factor distributions are in Supplementary Table 2. Since neither reference dataset has genetic information, we randomly assigned each individual in the reference data sets a normally distributed log PRS value with variance 1, mean 0 for individuals without a first-degree family history, and mean $\delta/2$ for individuals with a first-degree family history, where $\delta$ is the log RR per-SD difference in PRS. We assumed that genetic factors are independent of non-genetic factors except for family history.

#### Absolute risk of breast cancer by PRS and modifiable risk factors

The absolute risk of developing breast cancer for a woman of age *a* over the time interval *a + s* can be calculated as

$R_{a,a+s}= \int_{a}^{a+s} \lambda_{0}\left( t \right)\exp\left( \mathbf{Z}'\boldsymbol{\beta} \right)\exp\left( -\int_{a}^{t} [\lambda_{0}(u)exp(\mathbf{Z}'\boldsymbol{\beta})+m(u)]du \right)dt$	(a)

Formula (a) holds under the assumptions that the risk factors **Z** act in a multiplicative fashion on the baseline hazard function $\lambda_{0}\left( t \right)$. Here, $\boldsymbol{\beta}$ represents a vector of log RRs associated with each of the risk factors in **Z**. The baseline hazard is calculated by dividing the population average age-specific incidence rates by the average RR, where the average is taken over the risk factor distribution; risk factor distributions were calculated separately for women younger and older than fifty using the KNHANES data. Formula (a) accounts for competing risks due to mortality from other causes through the age-specific mortality rate function *m(t)*.

Cumulative risk is evaluated as absolute risk between age 20 years and a specific age. The 10-year risk is evaluated as absolute risk over the next 10 years for a woman who has attained a specific age without developing breast cancer. To use this method, we estimated the multivariable RRs of each woman in the KCPS-II based on their risk factors **Z**, the log RRs ($\boldsymbol{\beta}$) estimated in the KCPS, the age-specific mortality rates of breast cancer in Korea, and the risk factor distribution in KNHANES.

We classified individuals in the top 20% of the PRS distribution as high PRS (corresponding RR$\geq$1.33), those in the bottom 20% as low PRS (corresponding RR$\leq$0.75), and those in the middle category (>20^th^ to <80^th^ percentile) as middle PRS. Individuals above the median of modifiable risk score distribution were classified as an elevated modifiable risk and those below the median were considered as reduced modifiable risk group. Modifiable risk scores were generated using predicted log RRs based on BMI, oral contraceptive use, alcohol intake, and hormonal replacement therapy use. For each of these six PRS $\times$ modifiable risk groups, we calculated absolute risks for hypothetical women who have RR equal to the average RR in that group.

We performed bootstrapping to generate distributions of estimated lifetime (by age 80 years) absolute risk accounting for sampling uncertainty in risk factor RRs. We created 1,000 random samples of log RR parameters for *k* variables ($\hat{\beta}_{1i},\hat{\beta}_{2i}$,…$,\hat{\beta}_{ki}$, $i$=1,…,1000) from multivariate normal distribution with mean equal to the empirical log RRs ($\hat{\beta}_{1},\hat{\beta}_{2}$,…,$\hat{\beta}_{k}$) and variance equal to the squared standard errors of the estimates. For each of the 1,000 replicates, we calculated absolute risks for each of the six strata as above. The quantiles of this simulated distribution were used to estimate 95% confidence intervals.

Furthermore, we performed 5-year absolute risk projections by three birth cohorts (1920-1949, 1950-1969, 1970-1995 years) to examine birth cohort effects. For birth-cohort specific rates, age-specific rates were estimated using data on breast cancer mortality and incidence in Korea from 2000 to 2015. Since the age-specific data were partially available in some birth cohorts, the absolute risk projections were estimated using different age ranges across the birth cohorts (20-40, 35-60, and 55-75 years old).

**S.Table 1**. Questionnaire risk factor distributions in the validation cohort: Korean Cancer Prevention Study-II Biobank and Korean Cancer Prevention Study

| **Breast cancer risk factors** | **Korean Cancer Prevention Study-II Biobank** | | **Korean Cancer Prevention Study** | |
| --- | --- | --- | --- | --- |
|  | **Age <50, N=31,572** | **Age ≥50, N=9,459** | **Age <50, N=341,018** | **Age ≥50, N=102,609** |
| **Age at baseline, years** | | |  |  |
| Median (range) | 36 (20-49) | 56 (50-80) | 33 (21-49) | 58 (50-80) |
| **Age at menarche, years** | | |  |  |
| ≤10 | 145 (0.5) | 25 (0.3) | 544 (0.2) | 253 (0.2) |
| 11 | 721 (2.3) | 12 (0.1) | 9,296 (2.7) | 678 (0.7) |
| 12 | 3253 (10.3) | 143 (1.5) |  |  |
| 13 | 5861 (18.6) | 465 (4.9) | 126,152 (37.0) | 10,915 (10.6) |
| 14 | 6848 (21.7) | 1069 (11.3) |  |  |
| 15 | 6037 (19.1) | 1695 (17.9) | 162,993 (47.6) | 37,474 (36.5) |
| ≥16 | 3978 (12.6) | 4106 (43.4) |  |  |
| ≥17 |  |  | 42,033 (12.3) | 53,289 (51.9) |
| Missing | 4729 (15) | 1944 (20.6) |  |  |
| **Parity** | | |  |  |
| Nulliparous | 139 (0.4) | 13 (0.1) | 2,887 (2.1) | 137 (1.3) |
| 1 birth | 3449 (10.9) | 301 (3.2) | 53,051 (38.4) | 1,308 (12.1) |
| 2 births | 8836 (28) | 1933 (20.4) | 70,736 (51.1) | 3,202 (29.7) |
| 3 births | 1597 (5.1) | 2196 (23.2) | 11,653 (8.4) | 6,150 (57.0) |
| Missing | 17551 (55.6) | 5016 (53) | 202,691 | 91,812 |
| **Age at first birth, years (among parous women)** | | |  |  |
| <20 | 0 | 11 (0.1) | 823 (0.5) | 9,911 (10.9) |
| 20-24 | 111 (0.4) | 318 (3.4) | 34,262 (18.9) | 52,588 (57.6) |
| 25-29 | 600 (1.9) | 564 (6) | 123,269 (68.1) | 24,947 (27.3) |
| ≥30 | 146 (0.5) | 95 (1) | 22,708 (12.5) | 3,865 (4.2) |
| Missing | 30715 (97.3) | 8471 (89.6) | 159,956 | 11,298 |
| **OC use** | | |  |  |
| Never | 17948 (56.8) | 3817 (40.4) | 179,151 (91.7) | 6,807 (68.8) |
| Ever | 4765 (15.1) | 948 (10) | 16,116 (8.3) | 3,091 (31.2) |
| Missing | 8859 (28.1) | 4694 (49.6) | 145,751 | 92,711 |
| **HRT use (among women age ≥50)** | | |  |  |
| Never | - | 7957 (84.1) | - | 7,889 (86.2) |
| Ever | - | 1502 (15.9) | - | 1,262 (13.8) |
| Missing | - | 0 | - | 93,456 |
| **Type of HRT use (among current users age ≥50)** | | |  |  |
| Current C-type | NA | NA | NA | NA |
| Current E-type | NA | NA | NA | NA |
| Missing |  |  |  |  |
| **Age at menopause, years (among women age ≥50)** | | |  |  |
| <40 | - | 126 (1.3) | - | 3,026 (3.2) |
| 40-44 | - | 374 (4) | - | 9,101 (9.6) |
| 45-49 | - | 1590 (16.8) | - | 28,584 (30.2) |
| 50-54 | - | 3652 (38.6) | - | 43,268 (45.7) |
| ≥55 | - | 1005 (10.6) | - | 10,712 (11.3) |
| Missing | - | 2712 (28.7) | - | 7,918 |
| **Height, m** | | |  |  |
| Median (range) | 16.0 (13.0, 19.0) | 15.6 (13.1, 17.6) | 15.8 (13.1-19.0) | 15.3 (13.1-18.0) |
| **Body mass index, kg/m2** | | |  |  |
| <18.5 | 27762 (87.9) | 6042 (63.9) | 36,983 (10.8) | 64,412 (62.8) |
| ≥18.5 - <25 | 3336 (10.6) |  | 271,575 (79.6) |  |
| ≥25 - <30 | 428 (1.4) | 2995 (31.7) | 30,288 (8.9) | 34,631 (33.8) |
| ≥30 |  | 356 (3.8) | 2,172 (0.6) | 3,566 (3.5) |
| Missing | 46 (0.1) | 66 (0.7) | 0 | 0 |
| **Alcohol intake, g/day** | | |  |  |
| None | 10441 (33.1) | 6550 (69.2) | 277,046 (81.4) | 90,035 (88.6) |
| <5 | 8321 (26.4) | 1068 (11.3) | 50,297 (14.8) | 6,939 (6.8) |
| 5-14 | 5402 (17.1) | 597 (6.3) | 9,987 (2.9) | 2,797 (2.8) |
| 15-24 | 1389 (4.4) | 144 (1.5) | 2,116 (0.6) | 857 (0.8) |
| 25-34 | 487 (1.5) | 62 (0.7) | 265 (0.1) | 140 (0.1) |
| 35-44 | 352 (1.1) | 48 (0.5) | 462 (0.1) | 485 (0.5) |
| ≥45 | 645 (2) | 106 (1.1) | 352 (0.1) | 319 (0.3) |
| Missing | 4535 (14.4) | 884 (9.3) | 493 | 1,037 |
| **History of BBD** | | |  |  |
| No | 30071 (95.2) | 9303 (98.4) | 279,933 (82.1) | 97,900 (95.4) |
| Yes | 1360 (4.3) | 82 (0.9) | 61,085 (17.9) | 4,709 (4.6) |
| Missing | 141 (0.4) | 74 (0.8) | 0 | 0 |
| **Breast cancer family history in first degree** | | |  |  |
| No | 31043 (98.3) | 9329 (98.6) | 294,769 (86.4) | 85,831 (83.6) |
| Yes | 529 (1.7) | 130 (1.4) | 46,249 (13.6) | 16,778 (16.4) |
| Missing | 0 | 0 | 0 | 0 |

* Risk factor distributions are reported as n (%), unless otherwise specified. BBD = benign breast disease, C-type = estrogen and progestogen combined, E-type = estrogen-only, HRT = hormone replacement therapy, OC = oral contraceptive.

**S.Table 2.** Parameters used for the development of the breast cancer risk prediction model.

| Breast cancer risk factors | US | | | | | Korea | | | | |
| --- | --- | --- | --- | --- | --- | --- | --- | --- | --- | --- |
|  | **Population**  **distribution (%)** | | **Relative risk**  **of breast cancer** | | | **Population**  **distribution (%)** | | | **Relative risk**  **of breast cancer (95% CI)** | |
|  | **Age <50** | **Age ≥50** | | **Age <50** | **Age ≥50** | | **Age <50** | **Age ≥50** | **Age <50** | **Age ≥50** |
| Age at menarche, years |  |  | |  |  | |  |  |  |  |
| ≤10 | 15.0% | 15.2% | | 1.19 | 1.19 | | 1.5% | 0.1% | 1.27 (1.23 ,1.31) | 1.35 (1.25 ,1.46) |
| 11 | 18.5% | 18.1% | | 1.09 | 1.09 | | 5.5% | 0.3% | 1.13 (1.09 ,1.16) | 1.16 (1.08 ,1.25) |
| 12 | 22.8% | 22.2% | | 1.07 | 1.07 | | 15.5% | 2.4% | 1.13 (1.09 ,1.16) | 1.16 (1.08 ,1.25) |
| 13 | 21.4% | 21.6% | | Ref. | Ref. | | 23.1% | 6.8% | Ref. | Ref. |
| 14 | 13.7% | 14.1% | | 0.98 | 0.98 | | 24.1% | 12.5% | 1.00 (0.97 ,1.03) | 1.00 (0.93 ,1.08) |
| 15 | 6.1% | 6.1% | | 0.92 | 0.92 | | 16.7% | 16.7% | 0.89 (0.86 ,0.92) | 0.86 (0.80 ,0.93) |
| ≥16 | 2.6% | 2.6% | | 0.82 | 0.82 | | 13.7% | 61.2% | 0.79 (0.77 ,0.81) | 0.74 (0.69 ,0.80) |
| Age at menopause, years (among women age ≥50) | | | |  |  | |  |  |  |  |
| <40 | - | 21.3% | | - | 0.67 | | - | 4.7% | - | 0.57 (0.54 ,0.61) |
| 40-44 | - | 13.3% | | - | 0.73 | | - | 9.8% | - | 0.69 (0.64 ,0.74) |
| 45-49 | - | 22.7% | | - | 0.86 | | - | 33.8% | - | 0.83 (0.78 ,0.89) |
| 50-54 | - | 30.3% | | - | Ref. | | - | 41.7% | - | Ref. |
| ≥55 | - | 12.4% | | - | 1.12 | | - | 10.0% | - | 1.20 (1.12 ,1.29) |
| Parity |  |  | |  |  | |  |  |  |  |
| Nulliparous | 20.5% | 17.6% | | Ref. | Ref. | | 28.0% | 6.5% | Ref. | Ref. |
| 1 birth | 18.5% | 15.1% | | 0.87 | 0.87 | | 15.4% | 0.8% | 0.87 (0.69 ,1.10) | 0.79 (0.68 ,0.92) |
| 2 births | 32.2% | 33.7% | | 0.81 | 0.81 | | 44.5% | 34.5% | 0.76 (0.60 ,0.96) | 0.63 (0.54 ,0.73) |
| 3+ births | 28.9% | 33.6% | | 0.71 | 0.71 | | 12.1% | 58.2% | 0.66 (0.52 ,0.83) | 0.50 (0.43 ,0.58) |
| Age at first birth, years |  |  | |  |  | |  |  |  |  |
| <20 | 38.8% | 39.7% | | Ref. | Ref. | | 2.7% | 8.3% | Ref. | Ref. |
| 20-24 | 25.5% | 32.1% | | 1.01 | 1.01 | | 39.6% | 52.0% | 1.17 (0.86 ,1.58) | 1.11 (0.87 ,1.40) |
| 25-29 | 20.2% | 17.9% | | 1.11 | 1.11 | | 41.1% | 34.6% | 1.36 (1.01 ,1.85) | 1.22 (0.96 ,1.55) |
| ≥30 | 15.5% | 10.4% | | 1.24 | 1.24 | | 16.6% | 5.0% | 1.59 (1.17 ,2.16) | 1.35 (1.07 ,1.72) |
| OC use |  |  | |  |  | |  |  |  |  |
| Never | 14.3% | 19.1% | | Ref. | Ref. | | 91.7% | 77.1% | Ref. | Ref. |
| Ever | 85.7% | 80.9% | | 1.12 | 1.14 | | 8.3% | 22.9% | 1.25 (0.85 ,1.83) | 0.69 (0.50 ,0.96) |
| Current OC use (among women age <50) | |  | |  |  | |  |  |  |  |
| Former/Never | 86.9% | - | | Ref. |  | | 92.9% |  | Ref. |  |
| Current | 13.1% | - | | 1.19 | - | | 7.1% |  | 1.25 (0.85 ,1.83) | - |
| Body mass index, kg/m2 |  |  | |  |  | |  |  |  |  |
| <18.5 | 1.5% |  | | 1.28 | Ref. | | 9.2% | 62.0% | 0.98 (0.93 ,1.03) | Ref. |
| 18.5 - <25 | 41.0% | 35.1% | | Ref. | Ref. | | 69.5% |  | Ref. | Ref. |
| 25 - <30 | 30.0% | 31.9% | | 0.92 | 1.13 | | 17.3% | 33.2% | 1.02 (0.97 ,1.07) | 1.35 (1.22 ,1.49) |
| ≥30 | 27.6% | 33.1% | | 0.74 | 1.25 | | 4.0% | 4.8% | 1.04 (0.99 ,1.10) | 1.82 (1.65 ,2.01) |
| Height, m† |  |  | |  |  | |  |  |  |  |
| mean (SD) | 16.2 (0.6) | 16.2 (0.6) | | 1.17 | 1.17 | | 16.0 (0.5) | 15.4 (0.6) | 1.20 (1.15 ,1.27) | 1.24 (1.16 ,1.33) |
| Alcohol intake, g/day |  |  | |  |  | |  |  |  |  |
| None | 41.8% | 45.0% | | Ref. | Ref. | | 8.9% | 34.2% | Ref. | Ref. |
| <5 | 42.2% | 39.7% | | 1.01 | 1.01 | | 69.2% | 57.7% | 1.03 (0.98 ,1.08) | 1.01 (0.92 ,1.10) |
| 5-14 | 12.4% | 12.1% | | 1.03 | 1.03 | | 11.7% | 4.0% | 1.07 (1.01 ,1.12) | 1.01 (0.92 ,1.11) |
| 15-24 | 2.6% | 2.3% | | 1.13 | 1.13 | | 4.9% | 1.9% | 1.10 (1.05 ,1.15) | 1.02 (0.93 ,1.11) |
| 25-34 | 0.6% | 0.7% | | 1.21 | 1.21 | | 1.8% | 0.6% | 1.13 (1.08 ,1.19) | 1.02 (0.93 ,1.12) |
| 35-44 | 0.3% | 0.2% | | 1.32 | 1.32 | | 0.0% | 0.0% | 1.17 (1.12 ,1.23) | 1.03 (0.94 ,1.12) |
| ≥45 | 0.2% | 0.2% | | 1.46 | 1.46 | | 3.4% | 1.6% | 1.21 (1.15 ,1.27) | 1.03 (0.94 ,1.13) |
| History of BBD |  |  | |  |  | |  |  |  |  |
| No | 83.3% | 83.2% | | Ref. | Ref. | | 98.9%† | 99.4%† | Ref. | Ref. |
| Yes | 16.7% | 16.9% | | 1.68 | 1.51 | | 1.1%† | 0.6%† | 5.07 (4.85 ,5.30) | 13.91(12.31 ,15.73) |
| Family history of breast cancer | |  | |  |  | |  |  |  |  |
| No | 93.0% | 86.4% | | Ref. | Ref. | | 98.0%† | 98.8%† | Ref. | Ref. |
| Yes | 7.0% | 13.6% | | 2.50 | 1.60 | | 2.0%† | 1.2%† | 1.06 (1.00 ,1.13) | 1.18 (1.02 ,1.36) |
| HRT use (among women age ≥50) | |  | |  |  | |  |  |  |  |
| Never | - | 58.3% | | - | Ref. | | - | 84.2% | - | Ref. |
| Current |  | 9.9% | |  | 2.19 | | - | 15.8% |  | 0.98 (0.62 ,1.55) |
| Former | - | 31.9% | | - | 1.00 | | - |  | - |  |
| Type of HRT use (among women age ≥50) | |  | |  |  | |  |  |  |  |
| Current C-type | - | 94.3% | |  | Ref. | | - | 94.8%† | - | NA |
| Current E-type | - | 5.7% | |  | 0.72 | | - | 5.2%† | - | NA |
| PRS – log(OR) |  |  | |  |  | |  |  |  |  |
| PRS-11_ASN_ |  |  | | 0.32^1^ | 0.32^1^ | |  |  | 0.30^6^ | 0.30^6^ |
| PRS-42_ASN_ |  |  | | 0.40^2^ | 0.40^2^ | |  |  | 0.34^6^ | 0.34^6^ |
| PRS-136_EUR_ |  |  | | 0.54^3^ | 0.54^3^ | |  |  | 0.44^6^ | 0.44^6^ |
| PRS-209_EUR_ |  |  | | 0.48^4^ | 0.48^4^ | |  |  | 0.43^6^ | 0.43^6^ |
| PRS-CSx |  |  | | 0.48^5^ | 0.48^5^ | |  |  | 0.54^6^ | 0.54^6^ |

standard deviation = SD, BBD = benign breast disease, BMI = body mass index, C-type = estrogen and progestogen combined, E-type = estrogen-only, HRT = hormone replacement therapy, OC = oral contraceptive. †simulated prevalence. Korean relative risks were estimated using multivariable Cox proportional hazards in the KCPS; missing data for each factor were coded as a separate category (the "missing indicator method").

^1^per SD log(odds ratio) estimates calculated using the formula: $\sqrt{\sum_{i=1}^{n} \beta_{i}^{2}\times2\times q_{i}\times(1-q_{i})}$, where β_i_ (per-allele log(odds ratio) for SNP_i_) and q_i_ (minor allele frequency for SNP_i_) were derived from Ishigaki et al [2020]; ^2^per SD log(odds ratio) estimates calculated using the formula: $\sqrt{\sum_{i=1}^{n} \beta_{i}^{2}\times2\times q_{i}\times(1-q_{i})}$, where β_i_ and q_i_ were derived from Ho et al [2022]; ^3^Michailidou et al [2017]; ^4^Mavaddat et al [2019]; ^5^Ho et al [2022]; ^6^estimated from KCPS-II.

**S.Table 3.** Data sources and assumptions for each questionnaire and polygenic risk score (PRS) model

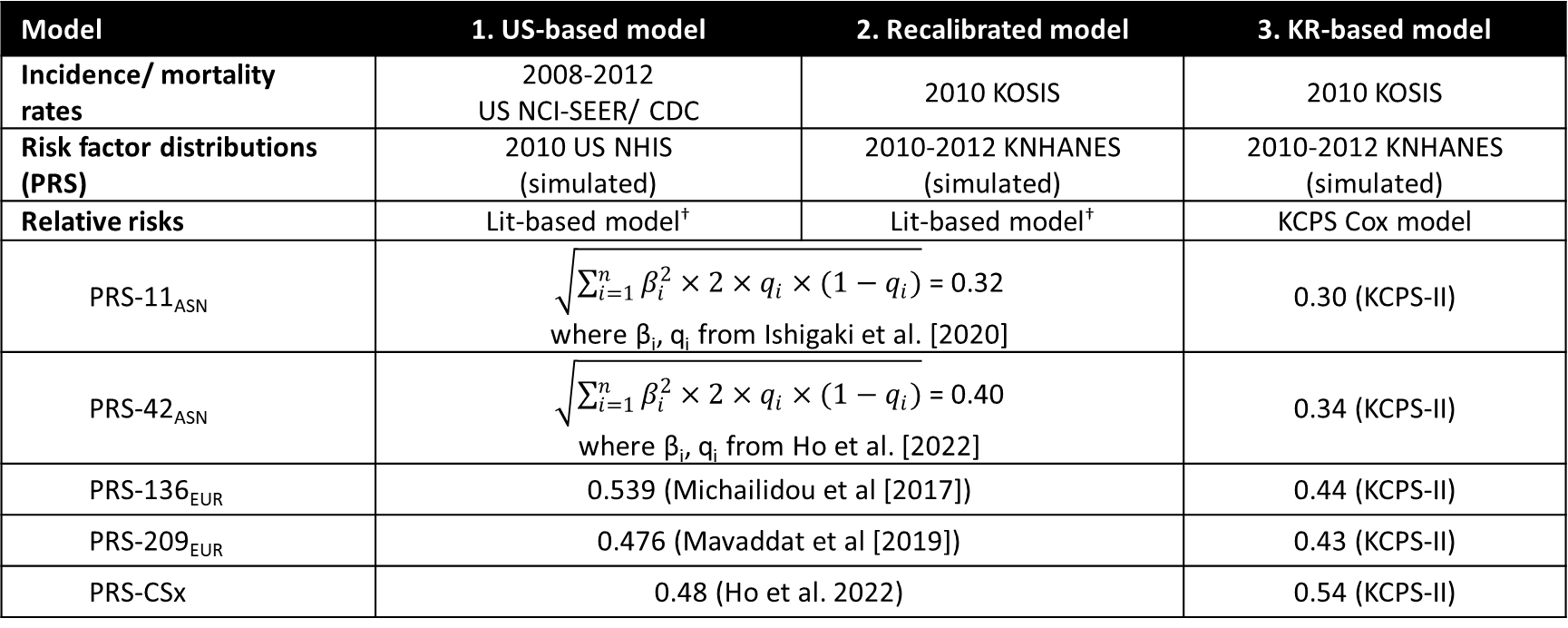

KOSIS = Korean Statistical Information Service; KNHANES = Korean NHANES; KCPS = Korean Cancer Prevention Study.

†Literature-based model: Garcia-Closas et al, JNCI 2014

**S.Table 4.** SNPs and beta coefficient SNPs used in the construction of PRS-11_ASN_, PRS-42_ASN_, PRS-136_EUR_, PRS-209_EUR_. (provided as an excel sheet)

**S.Table 5.** Discrimination and calibration of PRS-CSx for the breast cancer risk prediction models validated

|  |  | PRS-CSx | | | | |
| --- | --- | --- | --- | --- | --- | --- |
|  |  | Age <50 (459 cases, 57,269 controls*) | | | Age 50+ (226 cases, 22,059 controls*) | |
|  |  | AUC (95% CI) | | E/O (95% CI) | AUC (95% CI) | E/O (95% CI) |
| US-based European-ancestry | Qx | 0.66 (0.64, 0.69) | | 0.97 (0.88, 1.06) | 0.54 (0.5, 0.58) | 1.92 (1.69, 2.19) |
|  | PRS only | 0.72 (0.7, 0.74) | | 1.23 (1.12, 1.34) | 0.63 (0.59, 0.67) | 2.7 (2.37, 3.07) |
|  | Qx + PRS | 0.72 (0.7, 0.75) | | 0.99 (0.9, 1.08) | 0.63 (0.6, 0.67) | 1.94 (1.7, 2.2) |
| Recalibration | Qx | 0.65 (0.63, 0.68) | | 0.78 (0.71, 0.86) | 0.54 (0.5, 0.58) | 0.72 (0.63, 0.82) |
|  | PRS only | 0.72 (0.7, 0.75) | | 0.79 (0.72, 0.87) | 0.62 (0.58, 0.66) | 0.75 (0.66, 0.85) |
|  | Qx + PRS | 0.72 (0.7, 0.74) | | 0.78 (0.71, 0.85) | 0.63 (0.59, 0.67) | 0.72 (0.63, 0.82) |
| Korean-based | Qx | 0.65 (0.62, 0.67) | | 0.89 (0.81, 0.97) | 0.54 (0.51, 0.58) | 0.78 (0.68, 0.89) |
|  | PRS only | 0.72 (0.7, 0.75) | | 0.79 (0.72, 0.87) | 0.62 (0.59, 0.66) | 0.75 (0.66, 0.86) |
|  | Qx + PRS | 0.72 (0.69, 0.74) | | 0.88 (0.8, 0.96) | 0.63 (0.59, 0.66) | 0.8 (0.7, 0.91) |

*AUC and Expected/Observed (E/O) ratio estimates across first and second 5-years of follow-up periods were calculated using the fixed effects inverse variance weighting method, excluding women diagnosed with breast cancer or lost to follow-up in the first 5 years from the second 5 years of follow up.

**S.Table 6**. AUC (95% CI) of Asian and European PRS for the breast cancer risk prediction models validated using the KCPS-II Biobank

|  |  | PRS-11_ASN_ | PRS-42_ASN_ | PRS-136_EUR_ | PRS-209_EUR_ |
| --- | --- | --- | --- | --- | --- |
| Age <50 (459 cases, 57,269 controls*) | |  |  |  |  |
| US-based European-ancestry | Qx | 0.66 (0.64, 0.69) | 0.66 (0.64, 0.69) | 0.66 (0.64, 0.69) | 0.66 (0.64, 0.69) |
|  | PRS only | 0.67 (0.65, 0.7) | 0.68 (0.66, 0.7) | 0.70 (0.68, 0.72) | 0.70 (0.68, 0.72) |
|  | Qx + PRS | 0.68 (0.66, 0.7) | 0.69 (0.67, 0.71) | 0.71 (0.68, 0.73) | 0.71 (0.68, 0.73) |
| Recalibration | Qx | 0.65 (0.63, 0.68) | 0.65 (0.63, 0.68) | 0.65 (0.63, 0.68) | 0.65 (0.63, 0.68) |
|  | PRS only | 0.67 (0.65, 0.69) | 0.68 (0.66, 0.7) | 0.70 (0.68, 0.72) | 0.70 (0.68, 0.73) |
|  | Qx + PRS | 0.67 (0.65, 0.7) | 0.68 (0.66, 0.7) | 0.70 (0.67, 0.72) | 0.70 (0.68, 0.72) |
| Korean-based | Qx | 0.65 (0.62, 0.67) | 0.65 (0.62, 0.67) | 0.65 (0.62, 0.67) | 0.65 (0.62, 0.67) |
|  | PRS only | 0.67 (0.65, 0.69) | 0.68 (0.66, 0.7) | 0.70 (0.68, 0.72) | 0.70 (0.68, 0.73) |
|  | Qx + PRS | 0.66 (0.64, 0.68) | 0.68 (0.65, 0.7) | 0.69 (0.67, 0.71) | 0.69 (0.67, 0.72) |
| Age 50+ (226 cases, 22,059 controls*) | |  |  |  |  |
| US-based European-ancestry | Qx | 0.54 (0.5, 0.58) | 0.54 (0.5, 0.58) | 0.54 (0.5, 0.58) | 0.54 (0.5, 0.58) |
|  | PRS only | 0.57 (0.53, 0.61) | 0.60 (0.56, 0.64) | 0.62 (0.59, 0.66) | 0.61 (0.57, 0.65) |
|  | Qx + PRS | 0.58 (0.54, 0.61) | 0.61 (0.57, 0.64) | 0.63 (0.59, 0.67) | 0.62 (0.58, 0.65) |
| Recalibration | Qx | 0.54 (0.5, 0.58) | 0.54 (0.5, 0.58) | 0.54 (0.5, 0.58) | 0.54 (0.5, 0.58) |
|  | PRS only | 0.57 (0.53, 0.61) | 0.59 (0.55, 0.63) | 0.62 (0.58, 0.66) | 0.6 (0.56, 0.64) |
|  | Qx + PRS | 0.58 (0.54, 0.62) | 0.60 (0.56, 0.64) | 0.62 (0.58, 0.66) | 0.61 (0.57, 0.65) |
| Korean-based | Qx | 0.54 (0.51, 0.58) | 0.54 (0.51, 0.58) | 0.54 (0.51, 0.58) | 0.54 (0.51, 0.58) |
|  | PRS only | 0.57 (0.53, 0.61) | 0.59 (0.55, 0.63) | 0.61 (0.57, 0.65) | 0.60 (0.56, 0.64) |
|  | Qx + PRS | 0.58 (0.54, 0.62) | 0.59 (0.55, 0.63) | 0.61 (0.57, 0.65) | 0.60 (0.56, 0.64) |

*AUC estimates across first and second 5-years of follow-up periods were calculated using the fixed effects inverse variance weighting method, excluding women diagnosed with breast cancer or lost to follow-up in the first 5 years from the second 5 years of follow up.

**S.Table 7**. Expected/Observed ratio (95% CI) of Asian and European PRS for the breast cancer risk prediction models validated using the KCPS-II Biobank

|  |  | PRS-11_ASN_ | PRS-42_ASN_ | PRS-136_EUR_ | PRS-209_EUR_ |
| --- | --- | --- | --- | --- | --- |
| Age <50 (459 cases, 57,269 controls*) | |  |  |  |  |
| US-based European-ancestry | Qx | 0.97 (0.88, 1.06) | 0.97 (0.88, 1.06) | 0.97 (0.88, 1.06) | 0.97 (0.88, 1.06) |
|  | PRS only | 1.22 (1.12, 1.34) | 1.22 (1.12, 1.34) | 1.21 (1.11, 1.33) | 1.22 (1.11, 1.33) |
|  | Qx + PRS | 0.98 (0.89, 1.07) | 0.97 (0.89, 1.07) | 0.97 (0.88, 1.06) | 0.97 (0.89, 1.06) |
| Recalibration | Qx | 0.78 (0.71, 0.86) | 0.78 (0.71, 0.86) | 0.78 (0.71, 0.86) | 0.78 (0.71, 0.86) |
|  | PRS only | 0.79 (0.72, 0.87) | 0.79 (0.72, 0.87) | 0.79 (0.72, 0.86) | 0.79 (0.72, 0.86) |
|  | Qx + PRS | 0.77 (0.71, 0.85) | 0.78 (0.71, 0.85) | 0.77 (0.71, 0.85) | 0.77 (0.71, 0.85) |
| Korean-based | Qx | 0.89 (0.81, 0.97) | 0.89 (0.81, 0.97) | 0.89 (0.81, 0.97) | 0.89 (0.81, 0.97) |
|  | PRS only | 0.79 (0.72, 0.87) | 0.79 (0.72, 0.87) | 0.79 (0.72, 0.86) | 0.79 (0.72, 0.86) |
|  | Qx + PRS | 0.87 (0.8, 0.96) | 0.87 (0.79, 0.95) | 0.87 (0.79, 0.95) | 0.87 (0.8, 0.95) |
| Age 50+ (226 cases, 22,059 controls*) | |  |  |  |  |
| US-based European-ancestry | Qx | 1.92 (1.69, 2.19) | 1.92 (1.69, 2.19) | 1.92 (1.69, 2.19) | 1.92 (1.69, 2.19) |
|  | PRS only | 2.69 (2.36, 3.06) | 2.69 (2.36, 3.07) | 2.69 (2.36, 3.06) | 2.69 (2.36, 3.07) |
|  | Qx + PRS | 1.93 (1.69, 2.2) | 1.93 (1.69, 2.19) | 1.92 (1.69, 2.19) | 1.92 (1.69, 2.19) |
| Recalibration | Qx | 0.72 (0.63, 0.82) | 0.72 (0.63, 0.82) | 0.72 (0.63, 0.82) | 0.72 (0.63, 0.82) |
|  | PRS only | 0.75 (0.66, 0.85) | 0.75 (0.66, 0.86) | 0.75 (0.66, 0.86) | 0.75 (0.66, 0.86) |
|  | Qx + PRS | 0.71 (0.63, 0.81) | 0.72 (0.63, 0.82) | 0.72 (0.64, 0.82) | 0.72 (0.63, 0.82) |
| Korean-based | Qx | 0.78 (0.68, 0.89) | 0.78 (0.68, 0.89) | 0.78 (0.68, 0.89) | 0.78 (0.68, 0.89) |
|  | PRS only | 0.75 (0.66, 0.85) | 0.75 (0.66, 0.86) | 0.75 (0.66, 0.86) | 0.75 (0.66, 0.86) |
|  | Qx + PRS | 0.79 (0.69, 0.89) | 0.79 (0.69, 0.90) | 0.79 (0.69, 0.90) | 0.79 (0.70, 0.90) |

*Expected/Observed ratio estimates across first and second 5-years of follow-up periods were calculated using the fixed effects inverse variance weighting method, excluding women diagnosed with breast cancer or lost to follow-up in the first 5 years from the second 5 years of follow up.
**S.Table 8.** Lifetime (by age 80 years) absolute risk and 95% confidence intervals from 1000 bootstrap resamples.

|  | Absolute risk estimate | 2.5th percentile of bootstrap | 97.5th percentile of bootstrap |
| --- | --- | --- | --- |
| Low PRS / Reduced modifiable risk | 0.012 | 0.006 | 0.027 |
| Low PRS / Elevated modifiable risk | 0.025 | 0.013 | 0.056 |
| Middle PRS / Reduced modifiable risk | 0.024 | 0.007 | 0.033 |
| Middle PRS / Elevated modifiable risk | 0.053 | 0.027 | 0.119 |
| High PRS / Reduced modifiable risk | 0.049 | 0.016 | 0.067 |
| High PRS / Elevated modifiable risk | 0.103 | 0.034 | 0.142 |

**S.Table 9.** Checklist for studies developing or validations PRS (PRS-RS. Wand et al Nature 2021, <https://www.nature.com/articles/s41586-021-03243-6>) provided as an excel sheet.

**S.Figure 1.** Study design of validation cohort: Korean Cancer Prevention study-II (KCPS-II) Biobank

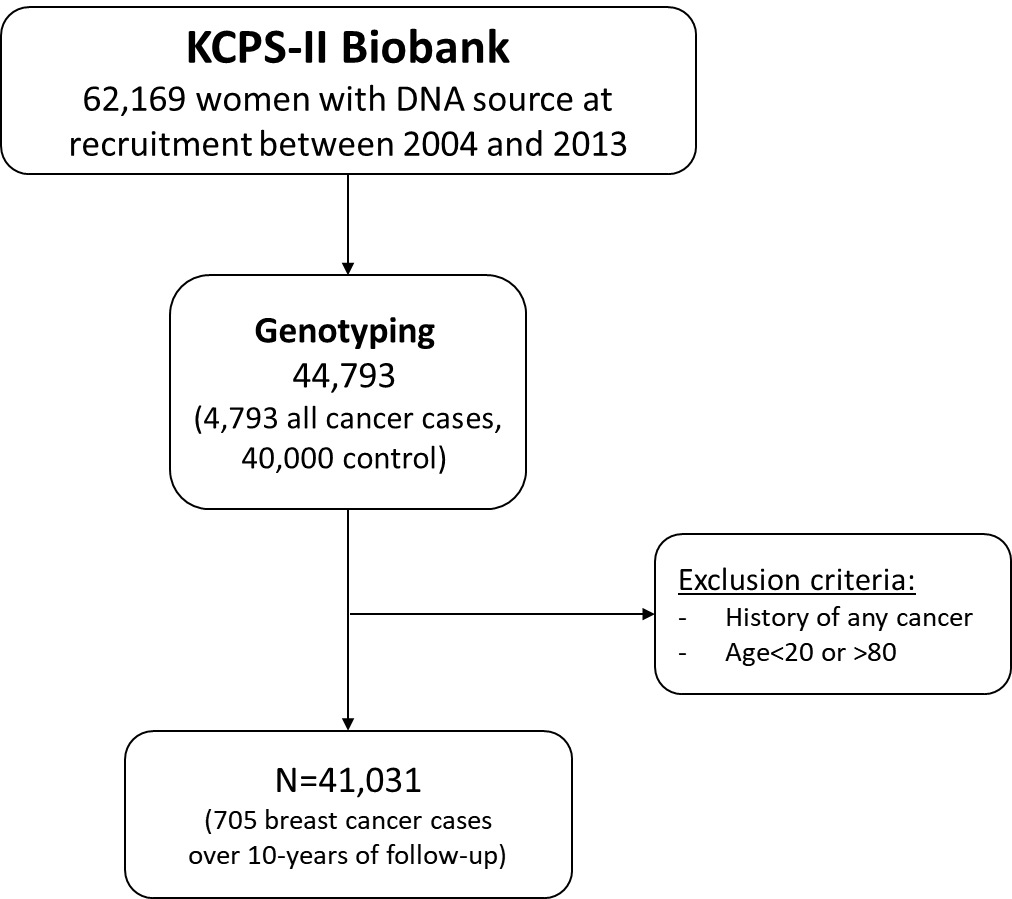

**S.Figure 2.** Overview of methods for PRSs development used in the study.

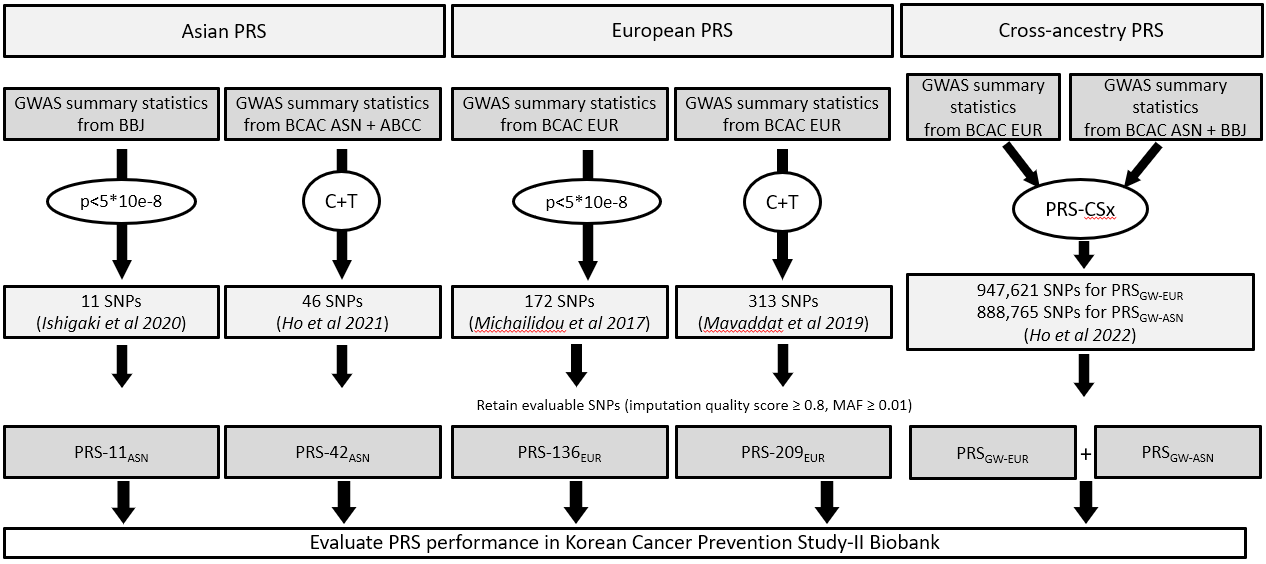

ABCC, Asia Breast Cancer Consortium; ASN, Asian; BBJ, The BioBank Japan Project; BCAC, Breast Cancer Association Consortium; C + T, clumping and thresholding; EUR, European; GWAS, genome-wide association study; PRS, polygenic risk score; SNP, single-nucleotide polymorphism.

**S.Figure 3.** Discrimination of Asian and European PRS for the breast cancer prediction models validated

| PRS-11_ASN_  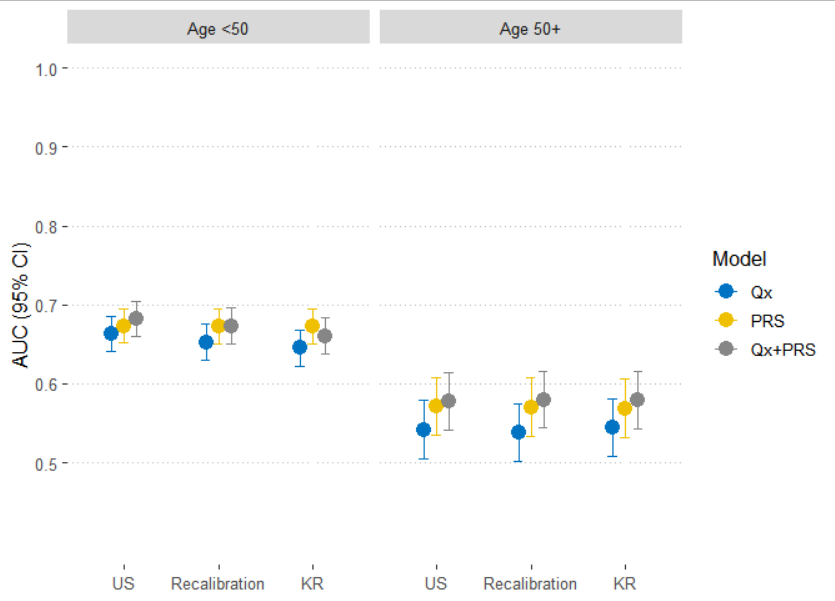 | PRS-136_EUR_  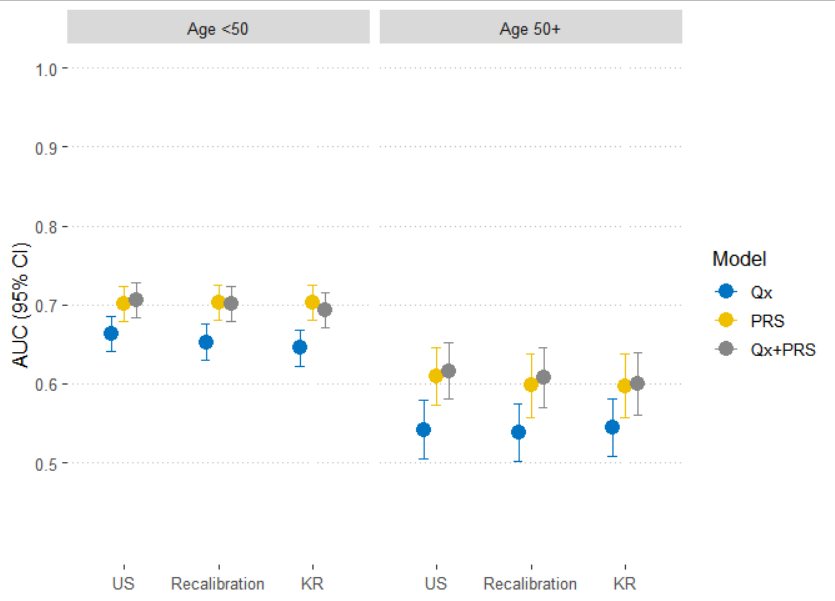 |
| --- | --- |
| PRS-42 _ASN_  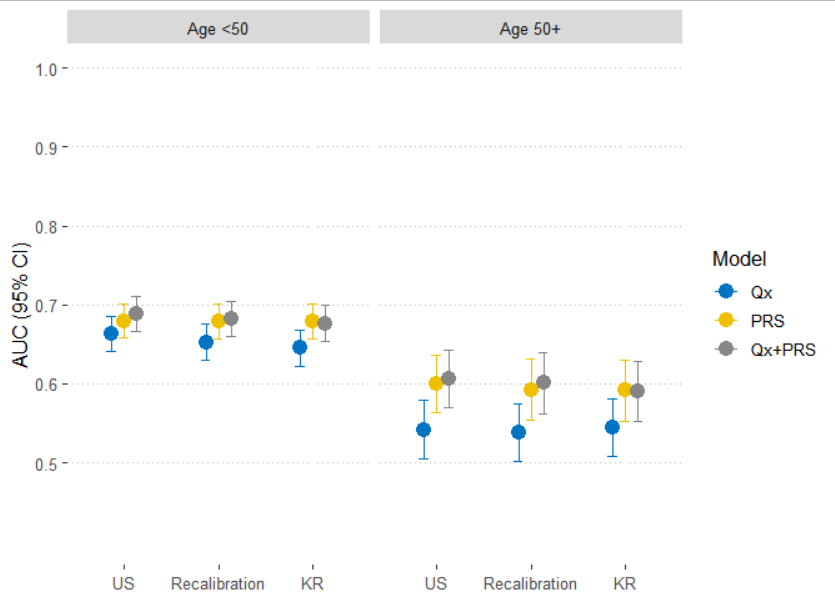 | PRS-209 _EUR_  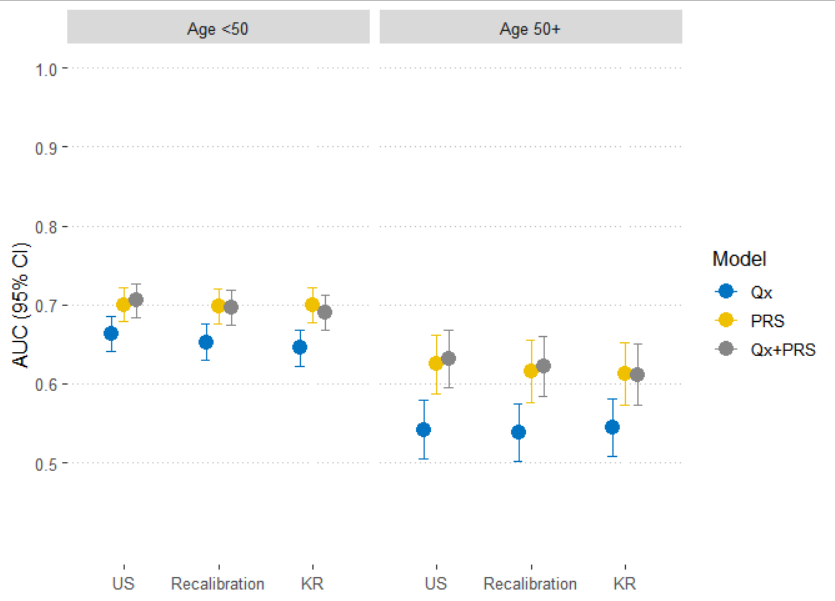 |

S.Figure 4. Calibration of Asian and European PRS for the breast cancer risk prediction models validated.

| PRS-11_ASN_  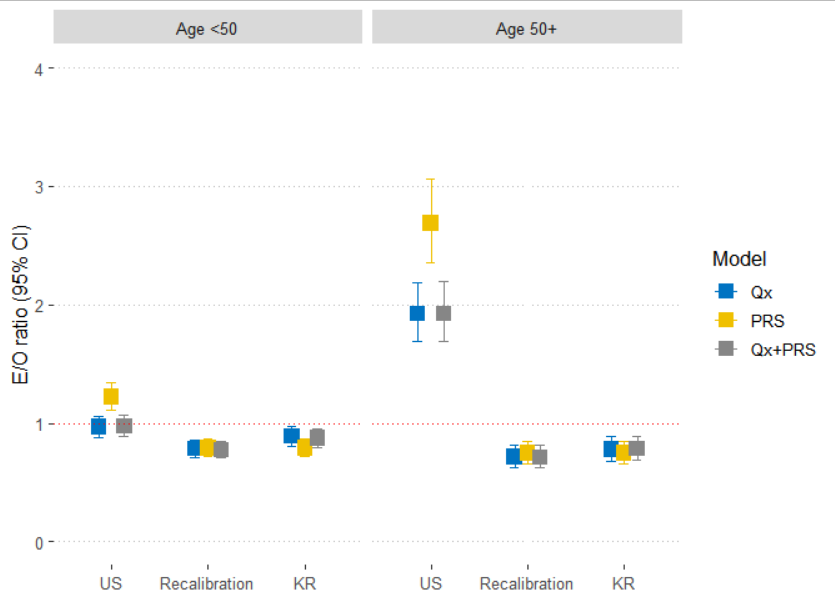 | PRS-136_EUR_  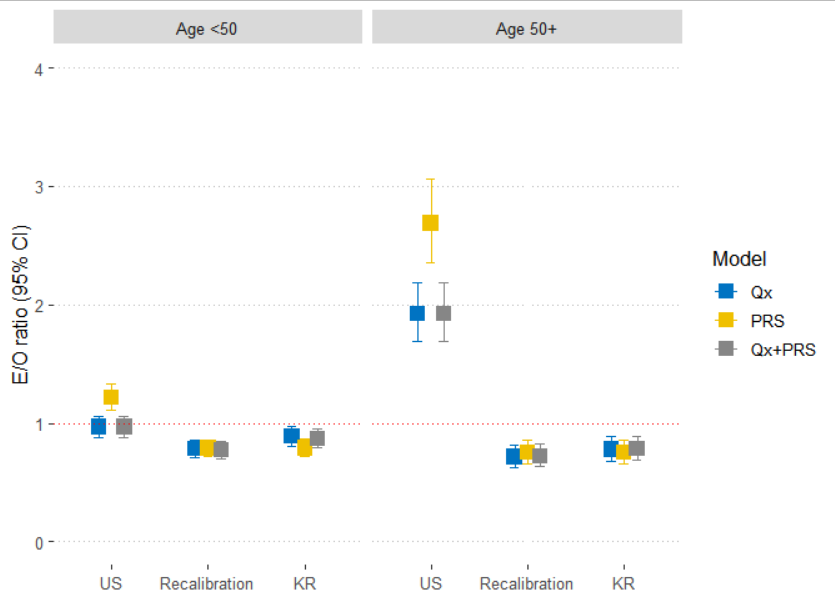 |
| --- | --- |
| PRS-42 _ASN_  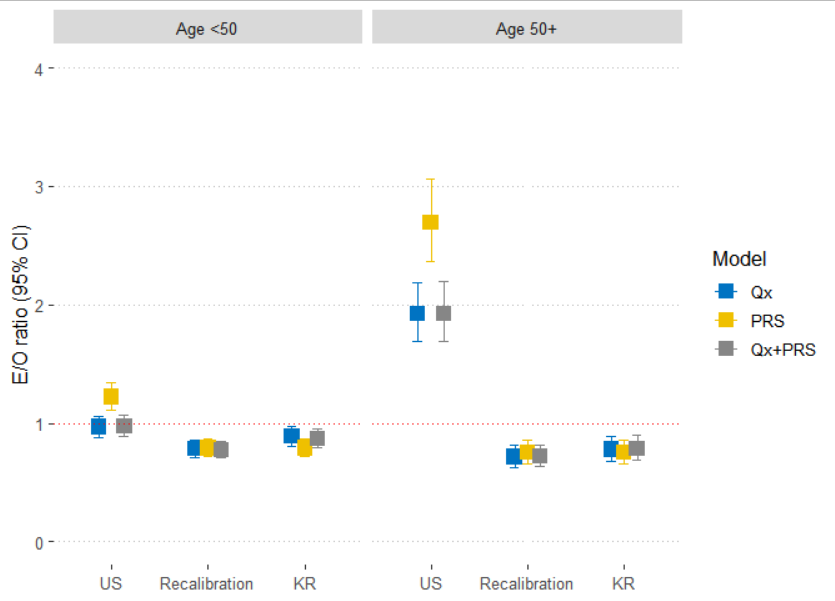 | PRS-209 _EUR_  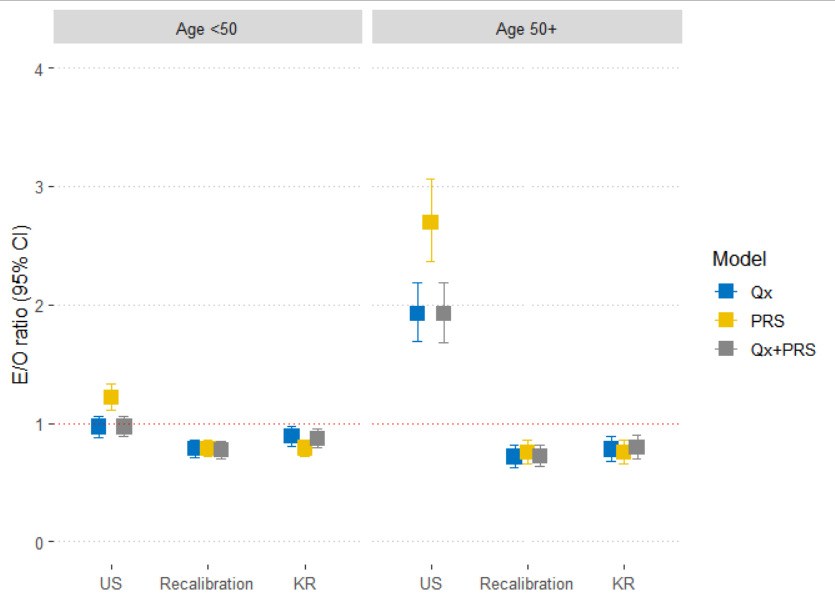 |
